# Barriers to access to antiretroviral therapy by people living with HIV in Indonesia during the COVID-19 pandemic: A qualitative study

**DOI:** 10.1101/2023.02.18.23285810

**Authors:** Nelsensius Klau Fauk, Hailay Abhra Gesesew, Alfonsa Liquory Seran, Paul Russell Ward

## Abstract

**Background:** The coronavirus disease (COVID-19) pandemic has a significant influence on access to healthcare services. This study aimed to understand the views and experiences of people living with HIV (PLHIV) about barriers to their access to antiretroviral therapy (ART) service in Belu district, Indonesia, during the COVID-19 pandemic.

**Methods:** This qualitative inquiry employed in-depth interviews to collect data from 21 participants who were recruited using a snowball sampling technique. Data analysis was guided by a thematic framework analysis.

**Results:** The findings showed that fear of contracting COVID-19 was a barrier that impeded participants’ access to ART service. Such fear was influenced by their awareness of their vulnerability to the infection, the possibility of unavoidable physical contact in public transport during a travelling to HIV clinic and the widespread COVID-19 infection in healthcare facilities. Lockdowns, COVID-19 restrictions and lack of information about the provision of ART service during the pandemic were also barriers that impeded their access to the service. Other barriers included the mandatory regulation for travellers to provide their COVID-19 vaccine certificate, financial difficulty, long-distance travel to the HIV clinic and a lack of public transport.

**Conclusions:** The findings indicate the need for dissemination of information about the provision of ART service during the pandemic and the benefits of COVID-19 vaccination for the health of PLHIV. The findings also indicate the need for new strategies to bring ART service closer to PLHIV during the pandemic such as a community-based delivery system. Future large-scale studies exploring views and experiences of PLHIV about barriers to their access to ART service during the COVID-19 pandemic and new intervention strategies are recommended.

## Background

Coronavirus disease, also commonly known as COVID-19, is a global public health problem. Since the report in Wuhan on December 31^st^, 2019 and the announcement by the World Health Organisation (WHO) in March 2020 as a global pandemic, COVID-19 has infected more than 580 million people and caused more than 6 million deaths worldwide as per the report on August 2^nd^, 2022 [1-3]. In the context of Indonesia, after the report of the first COVID-19 cases in March 2020, the number of cases reached 6,210,794 as per the report on August 2^nd^, 2022, and of whom 6,005,981 people were fully recovered, 47,809 people were hospitalised and 157,004 people died [4, 5]. COVID-19 has disrupted many aspects and sectors, including the economic, educational and health sectors. In the health sector, for example, COVID-19 has disrupted up to 75% of the provision of HIV care services and influenced the access of people living with HIV (PLHIV) to these services, especially antiretroviral therapy (ART) service [6-8].

Globally, reports and studies have predicted a range of possible factors or barriers that can influence or impede access of PLHIV to HIV treatment or ART service, which could lead to severe impacts including increased acquired immune deficiency syndrome (AIDS) related deaths [7, 9-11]. For example, a mathematical model study in Sub-Saharan Africa has suggested that the COVID-19 situation and transmission may limit the movement of PLHIV, which hinders their access to HIV care and treatment [7]. Similarly, several studies in different settings have reported a high possibility of interruption of HIV treatment or ART for PLHIV due to a reduction in the provision of the service and difficulty for PLHIV in accessing the service [12-16]. Shortage production of antiretroviral medicines by drug manufacturers is also reported by the WHO, which is predicted to also influence the availability and distribution of the medicines for PLHIV in many countries, especially the ones with poor resources [11]. Diversion of the focus of healthcare professionals, such as nurses and doctors, who care for PLHIV to care for COVID-19 patients is also considered to have an influence on the provision of care for PLHIV in many settings throughout the world [15, 17-19]. In addition, some previous findings have suggested that lockdowns and interstate travel bans have played a role as barriers that hampered PLHIV in some settings from accessing ART or refilling their medication [9, 20-22]. Other influencing factors for the access of PLHIV to ART are fear of contracting COVID-19, perceived increased risk of acquiring COVID-19 when coming to healthcare clinics, confusing COVID-19 restrictions, abuse by police and soldiers at roadblocks and shortage of personal protective equipment [23, 24].

The existing literature suggests that there is still limited evidence on how COVID-19 has impacted the access of PLHIV to HIV treatment or ART service globally. Similarly, in the context of Indonesia, there have never been studies exploring the influence of COVID-19 on the access of PLHIV to ART service. Therefore, this study aims to fill in this gap in knowledge by exploring the views and experiences of PLHIV in Belu district, Indonesia about barriers to their access to ART service during the COVID-19 pandemic. Understanding these barriers can be useful to develop new strategies that can address the needs of PLHIV and help bring HIV care and treatment service closer to them during the COVID-19 pandemic.

## Methods

### Study setting

The study was carried out in Belu district which is located in the eastern part of Indonesia and shares the border with East Timor [25]. Belu covers an area of 1,284,94 km^2^ occupied by a total population of 204,541 people [25, 26]. The district has 12 sub-districts, 17 public health centres and four hospitals (one public hospital, one army hospital and two private hospitals) [26, 27]. COVID-19 healthcare services during the pandemic are only provided in the public hospital and a private hospital due to limited COVID-19 trained healthcare professionals and medical devices. The district has one HIV clinic where ART service is provided for more than a thousand PLHIV.

### Recruitment procedure and data collection

This study employed a qualitative design to collect data from 21 PLHIV. It aimed to understand the views and experiences of people living with HIV (PLHIV) about barriers to their access to antiretroviral therapy (ART) service in Belu district, Indonesia, during the COVID-19 pandemic. Participants were recruited using exponential non-discriminative snowball sampling technique through which initial participants were asked for help to provide multiple referrals or distribute the study information sheets to their eligible friends and colleagues who might be willing to take part in the study [28]. Initially, the study information sheets were distributed to potential participants through six public health centres (Pusat Kesehatan Masyarakat) and through nurses (who were also HIV counsellors) who run HIV services (HIV testing and counselling) at those public health centres. Potential participants who contacted the field researcher (ALS, female) to confirm their participation were recruited for an interview. The researcher is a pharmacist, has attended formal training in qualitative methods and regularly conducted qualitative studies on different public health issues. Some potential participants verbally stated their willingness to participate to those nurses at public health centres as they were not possessing a mobile phone to directly contact the field researcher. The nurses directly linked these potential participants to the field researcher via phone calls and an agreement to meet for an interview was directly discussed between the researcher and each of the potential participants. This snowball sampling technique helped the distribution of the study information among PLHIV who were currently accessing ART, stopping ART due to the COVID-19 pandemic or just restarting ART when this study was conducted. Recruitment of the participants was based on several inclusion criteria, including one had to be HIV-positive, aged 18 years old or above, and currently access or had previously accessed ART service prior to the COVID-19 pandemic. Finally, twenty-one PLHIV were recruited and participated in this study. None of the participants who had confirmed to participate in this study withdrew their participation.

The participants were male (n=14) and female (n=7) living with HIV. Their age ranged from 25 to 53 years old, with the majority in the group age 31 to 40 years old (n=12), followed by the age group 41 to 50 (n=6). Most participants graduated from high school (n=13), one had a diploma or bachelor’s degree and the rest graduated from elementary school or were school dropouts. All the female participants were housewives, while the male participants engaged in different kinds of professions, with the majority having professions as motorbike taxi drivers and construction workers (n=9) and the others were fishermen (n=2), school teacher (n=1) and unemployed (n=2).

Data collection was conducted using one-on-one and face-to-face in-depth interviews from April to May 2022. Interviews were carried out in a private room at the public health centres where the participants accessed general healthcare services. The time and venues for interviews were mutually agreed upon by the field researcher and the participants. Interviews were focused on exploring participants’ views and experiences about barriers that influenced or impeded their access to ART service during the COVID-19 pandemic. These included the participants’ views and feeling about the COVID-19 pandemic; their views and experiences of interacting with other people during the pandemic; their views about the lockdown, COVID-19 mandatory protocols or restrictions and vaccine and how these factors influenced their activities; the impact of COVID-19 pandemic and its mandatory protocols on their work and income; their views about their health condition in relation to the widespread COVID-19 infection; and how these views and experiences influenced their access to HIV treatment or ART service during the pandemic. Interviews took 35-50 minutes and were conducted in Indonesian, the primary language of the field researcher and the participants. Interviews were recorded using an audio digital recorder and the interviewer sometimes took notes when she felt necessary. Only the interviewer and each participant were present in the interview room during the interviews and both maintained physical distancing of 1.5 metres and wore a facemask. None of the participants had established relationships with the researchers prior to the interviews. Interviews and the recruitment of the participants ceased when the researchers felt that information or data provided by the participants had been rich enough to address the research questions and objective. This was determined when data saturation had been reached which was indicated in the similarities of responses provided by the last few participants to those of previous participants. No repeated interviews were conducted with any of the participants. The participants were offered an opportunity to read and provide comments or corrections, after transcription, to the information they provided in the interviews, but none of them took the offer.

### Data analysis and ethical consideration

The comprehensive data analysis was conducted after the verbatim transcription of audio recordings into coding sheets. The comprehensive data analysis was conducted in Indonesian to minimise the risk of losing meanings attached to idiomatic expressions [29]. For the purposes of publication, the selected quotes were translated into English by the first author (NKF) who is fluent in both Indonesian and English. The translations were then checked for accuracy and clarity by other authors (ALS, HAG, PRW). Data analysis was performed manually and guided by five steps of qualitative data analysis introduced in Ritchie and Spencer’s qualitative thematic framework analysis [30, 31]. These five steps were (i) familiarisation with the data which was performed by reading each transcript repeatedly, breaking down information in the transcript into chunks of information, highlighting information, providing comments and labels to chunks of information to look for ideas and patterns; (ii) identification of a thematic framework by listing the emerging issues, concepts and ideas in each transcript and these were then used to form the thematic framework; (iii) data were indexed by creating open codes to chunks of information in each transcript. This was followed by creating close coding through which similar or redundant codes were identified and then collated to reduce the number of codes. The codes were then grouped thematically; (iv) data were charted by reorganising and rearranging the themes and the codes in a summary chart to enable comparison of data within each transcript or across transcripts; and (v) finally, mapping and interpretation of the entire data [31-33].

This study obtained its ethics approval from the Health Research Ethics Committee, Duta Wacana Christian University, Indonesia [No. 1005/C.16/FK] and all the methods were performed in accordance with the guidelines of the Declaration of Helsinki. Participants received information about the study through the study information sheets distributed to them during the recruitment process. They again received verbal and in-person explanations from the field researcher about the study’s objective and the use of the data and were allowed to raise any concerns or ask for clarification before the interviews. They were informed that their participation was voluntary and that they had the right to withdraw from the study at any time without any consequences. They were also assured that the confidentiality of their identity during the recruitment process and after the interviews and the anonymity of data or information they provided will be maintained to prevent the possibility of the link of data or information to any individuals in the future. This was done by providing pseudonyms to each transcript for the de-identification purpose. Before commencing the interviews, each participant signed and return an informed consent form.

## Results

The analysis resulted in four main emerging themes about COVID-19-related barriers to access to ART by PLHIV during the pandemic. They were (i) fear of COVID-19 transmission (e.g., in public transport during travel to the HIV clinic or in healthcare facilities), (ii) lockdown and lack of information about ART service delivery during the COVID-19 outbreak, (iii) mandatory COVID-19 vaccine certificate for travellers, and (iv) financial barriers. Detailed analysis of these themes is presented below.

### Fear of COVID-19 transmission

COVID-19 pandemic and its severe impacts including deaths have created fear among group and community members including PLHIV, which influenced how people engaged in their activity and work and interacted with each other. For example, fear of contracting COVID-19 infection was a factor that influenced the access of PLIHIV, both female and male, to HIV treatment or ART service during the pandemic. Marni, a woman in who had just restarted ART a month prior to the study, explained:

> *“When COVID-19 cases were rampant, it was really scary, I mean it is still scary until now. I did not travel to the [HIV] clinic to collect the [antiretroviral] medicines, because I was afraid of getting corona. I am still scared up to now”*.

Participants’ fear of COVID-19 transmission which hampered their access to ART service seemed to be supported by their awareness of how vulnerable they were to the infection and the possibility of negative impacts they could experience if they had COVID-19 and HIV co-infections. The following narrative of Joni who had been living with HIV for years, reflects such fear and perception:

> *“I didn’t want to go anywhere during those months when the corona cases were high because I was afraid of contracting the virus. I am aware that my body is already infected with HIV and if I get infected with COVID-19 as well, then the impact on my health could be very bad. I think I am definitely more susceptible to COVID-19 than people who are HIV-negative. I am still scared of having HIV and COVID co-infections. That is why I decided not to go anywhere far from home including not going to the HIV clinic to collect the [antiretroviral] medicines, and I decided to wait until the condition gets better. I plan to go back to access the medicines next month if possible”*.

Joni’s narrative, which was also echoed by other participants, reflected a lack of consideration of the possible detrimental effects of interruption of ART on their health, which may result in viral rebound, immune decomposition or clinical progression. Such a lack of consideration might be stemming from a limited knowledge about these negative effects, and this has important implications for healthcare professionals and systems to ensure that PLHIV are aware of the consequences of ART discontinuation.

The fear among the participants was also influenced by the poor transportation system which was reflected in the limited availability of public transport. Narratives in this study showed that some participants were fully aware of the poor transportation system which would lead to them meeting, interacting and having physical contact with other people during a travelling to the HIV clinic which could increase the possibility of COVID-19 transmission to them. Such awareness seemed to be supported by the fact that the minibuses (known as mikrolet) or public transport in the study setting were not designed to support physical distancing, which made physical contact unavoidable. Similarly, participants were also aware of the high possibility for them to meet other people on the street and in the hospital where the HIV clinic is located, which was considered to increase their chance to catch the virus:

> *“The COVID-19 situation has been very scary for me. You can imagine if one of our family members dies due to COVID-19 then we can’t even be close to him or her to see their face, other people [COVID-19 officers] are the ones who take care of the body and funeral which is in certain locations that are far away, very sad. It also creates tremendous fear. That’s why I thought that it’s better if I don’t have to travel far to the HIV clinic to access to ART service. You can imagine I have to go by public transport such as a minibus where passengers sit close to each and there is no distance. The possibility of contracting the coronavirus is very high. Moreover, I will certainly meet a lot of people on the minibus or the street or in the hospital, I could contract the virus. That’s why I decided to postpone access to the medicines” (Sefa, female)*.

Public perception that COVID-19 was widespread in hospitals and other healthcare facilities where COVID-19 patients were treated was another factor that influenced both female and male participants’ access to ART service at the HIV clinic which is a part (ward) of a public hospital in the study setting. A female stated:

> *“Public hospitals are where corona spreads, everyone is afraid to go to public hospitals because many people who have the virus are treated there*.*”*

Such perception was underpinned by the information they received from healthcare professionals who encouraged their family members to avoid an unnecessary visit to two hospitals where COVID-19 patients were treated:

> *“I myself decided not to go to collect the [antiretroviral] medicines at the public hospital [where the HIV clinic is located] because there are a lot of COVID-19 patients there. I heard that one of the nurses who work for the public hospital said that there are many COVID-19 patients in the public hospital, so unnecessary visits to the hospital should be avoided. A health worker said this, so it must be true, that’s why I didn’t go there [the hospital where the HIV clinic is located] for several months because I was afraid of getting COVID” (Anis, male)*.

### Lockdown and lack of information about ART service delivery during the COVID-19 outbreak

The enforcement of COVID-19 mandatory lockdown and restrictions on community activities that involved physical contact due to the escalation of COVID-19 cases was reported as another barrier that impeded both female and male participants’ access to HIV treatment or ART service. Tinus, a male participant, explained:

> *“During the lockdowns, I did not go to the clinic to get my medicines, I did not work either. I mostly just stayed at home”*.

The findings also pointed to the poor healthcare system which was reflected in the limited dissemination of information and the lack of understanding among participants about ART service provision during the COVID-19 pandemic. These seemed to lead to the uncertainty and reluctant among participants about whether or not to access the service, as illustrated in the following story of a woman who has been living with HIV for more than 10 years:

> *“I didn’t access the [antiretroviral] medicines for several months during the lockdowns. There were announcements by the government about the lockdowns, and I knew that it was not allowed to visit crowded places and that people needed to stay at home to avoid the transmission of COVID-19. So, I was not going anywhere, especially when the number of COVID-19 cases was rising sharply and people died from COVID nearly every day. There wasn’t any information about the provision of the [ART] service during COVID. I lived in the village so what I knew was that there were restrictions to go to crowded places and people need to maintain physical distancing…*..*” (Shinta)*.

Similarly, participants’ lack of understanding of COVID-19-related lockdowns and restrictions, which seemed to be stemming from limited dissemination of information within communities by the government, had an influence on some participants’ access to ART service during the pandemic. For example, the perception that lockdowns meant no healthcare services for people or patients other than COVID-19 patients was a misunderstanding that hindered some participants’ access to ART:

> *“I didn’t know whether the HIV clinic was open or not during the lockdowns. People said that lockdown means everything is closed, and patients are not served except COVID-19 patients, so I didn’t go to the clinic. I lived very far from the clinic, if I went there and it turned out that the clinic was closed, then I spent money on transportation for nothing. So, at that time I thought that I should wait until the COVID-19 situation gets better. I have restarted the therapy” (Lukas, male)*.

### Mandatory COVID-19 vaccine certificate for travellers

Mandatory regulation for inter-district travellers to bring and provide their COVID-19 vaccine certificate to the police or municipal police was also a barrier that impeded some female and male participants’ access to ART service during the COVID-19 outbreak. Participants described how municipal police and police were placed on standby for checking travellers’ COVID-19 vaccine status and certificate, which often resulted in unvaccinated travellers being denied to enter another district. Such seemed to discourage some unvaccinated participants to access ART service for months when COVID-19 mandatory lockdown protocols were put in place:

> *“At the time when the number of COVID-19 cases increased rapidly and people died from COVID-19 nearly every day, I heard that municipal police and police were placed on standby at many spots and I knew that I was not vaccinated so I thought they wouldn’t allow me to come to XX [name of the place] even if I provide them with my identity card. That was the reason why I didn’t collect the [antiretroviral] medicines. …. At that time I was in XXX [another district] for several months. I have family here [name of the place] and also over there [name of the place]” (Beni, male)*.
>
> *“I moved to XXX (name of the district) for more than a year, when there were many COVID cases in XXX [name of the district]. I stayed there with my parents. I live here [name of the place] but originally come from XXX [name of the place]. I didn’t come to collect my [antiretroviral] medicines for months because I was in XXX [name of the place], not vaccinated, and didn’t have a vaccine certificate” (Ani, female)*.

Some participants who had not received the COVID-19 vaccine also raised their concerns about the possibility of negative effects of the vaccine on their health due to their HIV-positive status. Such concerns are reflected in statement like:

> *I have not been vaccinated because I am afraid that the COVID-19 vaccine might worsen my health condition because I am living with HIV’ (Yoseph, male)*.

Their concerns about the COVID-19 vaccine seemed to be influenced by a lack of knowledge about the COVID-19 vaccine and misleading information about the negative side effects of the vaccine as illustrated in the following narrative:

> *“I have not been vaccinated because I am afraid of any negative effects on my health. Now everyone is suddenly being told to receive the COVID-19 vaccine but they don’t know what the negative effects will be, especially for people with HIV like me. I heard some people said that the COVID-19 vaccine could have negative impacts on people living with HIV like me, that’s why I have doubts and haven’t received the vaccine” (Ima, female)*.

### Financial barriers

Poor economic or financial condition due to the disruption of jobs was a negative impact COVID-19 pandemic experienced by both male and female participants, which was considered as an influencing factor for their access to ART service. Participants described how the COVID-19 situation had affected their income generating activities and led to the experience of significant income reduction or loss of income. These put them into financial hardships which also influenced their access to ART service due to the inability to afford the related costs. The following narratives of a female and male participant reflect the influence of the COVID-19 outbreak on their financial condition which negatively impacted their life in general and their access to HIV treatment in particular:

> *“Before the COVID-19 outbreak, I was selling cakes like fried bananas in an elementary school complex, so my income was pretty good every day. However, when COVID-19 occurred and the number of COVID cases increased, I was prohibited from selling cakes at the school because there was a concern about the possibility of spreading COVID-19 to students. Since then, I immediately lost my income and had no income at all. I am having financial difficulties to meet the needs of my family, and this is also the reason why I don’t go to the HIV clinic to collect the (antiretroviral) medicines, I don’t have money for transport and administration…” (Yustin, female)*.
>
> *“COVID-19 pandemic ruins my income until now, it seems that people are reluctant to go by motorcycle taxi due to the fear of contracting COVID-19. Many motorcycle taxis, including myself, have significant income reduction during this pandemic. It is very difficult to earn money during this pandemic. So, I didn’t go to XXX [place where the HIV clinic is located] to collect the [antiretroviral] medicines because I had to spend money on gasoline and also food and drinks. It takes nearly an entire day to go to XXX [name of the place] and come back here. But I went back to collect the medicines last month” (Benyamin, male)*.

Financial consequences were also experienced by participants due to long-distance travel to the HIV clinic, which required them to spend more money on transport. This was acknowledged as hindering their access to ART service as illustrated in the following story:

> *“The distance from here to XXX [place where the HIV clinic is located] is far and there is no minibus [public transport]. So, if I want to go to the HIV clinic, then I have to use a motorcycle taxi and the cost is quite expensive because of the long distance. The problem is that I am financially dependent on my parents, and during this time of COVID-19 they have no income, so I don’t go to the clinic to collect the [antiretroviral] medicine” (Petrus, male)*.

The above narratives indicated that difficult economic or financial conditions experienced by the participants may also have an influence on their ability to afford costs for opportunistic infection treatment prescriptions or other health issues they and their family members experienced. In addition, such a difficult condition seemed to lead them to making difficult decisions on allocating the budget for basic necessities over spending it for costs related to accessing ART or other health care services.

## Discussion

The COVID-19 pandemic is reported to have unprecedented detrimental impacts on HIV care services and PLHIV as it has caused the disruption of ART in many countries, influenced the access of PLHIV to the therapy, worsened clinical outcomes and increased mortality among PLHIV across the globe [7, 9-11, 34, 35]. This study explored views and experiences of PLHIV in Belu district, a resource-limited setting in Indonesia, about barriers to their access to ART service during the COVID-19 pandemic. Overall, it highlights several barriers to ART service access among PLHIV, including fear of contracting COVID-19 transmission, lockdown and lack of information about ART service delivery during the COVID-19 pandemic, mandatory COVID-19 vaccine certificate for inter-district travellers and financial barriers.

The fear of contracting COVID-19 infection, which influenced their access to ART service, was amplified by the awareness of certain conditions that can increase the risk of transmission, such as unavoidable physical contact with other people in public transport to the HIV clinic and the widespread COVID-19 infection in the public hospital, where the clinic is located. The findings support the report of a previous study suggesting the perceived increased risk of contracting COVID-19 infection in healthcare facilities as an influencing factor for the access of PLHIV to ART service during the pandemic [23]. Our findings also indicate a bigger picture of poor public transportation and healthcare systems in the study setting. These are reflected in a lack of public transport and the unavailability of ART service within communities, which seemed to contribute to the various challenges facing PLHIV. The current study shows that participants’ fear and awareness of how vulnerable they are to COVID-19 infection and the high possibility of COVID-19/ HIV co-infections which can lead to severe negative health impacts also hampered their visit and access to ART service. The participants’ fear of COVID-19/HIV co-infections and the negative impact is plausible as previous studies have reported evidence of increased risk for COVID-19 infection progression or worse clinical outcomes and increased mortality among PLHIV who are infected with COVID-19 [10, 34, 36, 37]. These findings are also in line with the concepts of perceived susceptibility and severity, which refer to the participants’ belief about the possibility of contracting COVID-19 and the seriousness of its impact, as supporting factors for performing recommended preventive behaviours (i.e., in this study: avoiding physical contacts in public transport, avoiding exposure spots such as healthcare facilities) [38, 39]. However, discontinuation of ART does not seem to be a positive option, is not recommended and may reduce the immunological benefits of treatment and increase complications related to HIV due to viral rebound and immune decomposition [40, 41]. The findings have important implications for the local health departments and healthcare systems in the study setting and other parts of Indonesia to bring ART service closer to PLHIV through public/sub-public health centres at sub-district and village levels or through a community-based model of ART delivery which has been proven effective in other settings or countries [42, 43]. These delivery strategies may help reduce the fear of contracting COVID-19 infection that prevents access to ART among PLHIV or increase their access to ART.

Consistent with previous findings [9, 20-22], this study confirms that lockdown and COVID-19-related restrictions during the escalation of COVID-19 cases were barriers that hindered participants’ access to ART service during the pandemic. In addition, this study adds further evidence suggesting that participants’ lack of information and misunderstanding about lockdowns and the restriction guidelines were also factors that influenced their access to the service during the pandemic. The findings reflect the unpreparedness of the healthcare system in the study setting to get PLHIV informed about both the healthcare service delivery, especially ART service during the pandemic, and the correct information about the COVID-19 pandemic’s mandatory lockdown guidelines. In other words, the current findings indicate less approachability of ART service or scarcity of the dissemination of healthcare-related information for PLHIV and other community members during the COVID-19 pandemic, which influenced their access to the service [44-48]. Approachability or well-known information about healthcare services within communities is an important healthcare service dimension that determines the accessibility of the services [44, 45]. As a consequence, there seems to be a low level of access to care-related health literacy among the participants which is an important aspect that can enable them to make critical decisions to seek and access appropriate healthcare services, especially ART during the COVID-19 pandemic [49, 50].

Our findings suggest that the requirement for inter-district travellers to provide a COVID-19 vaccine certificate was another identified factor that impeded some unvaccinated participants’ access to ART service during the pandemic. This adds further evidence to the previous findings which have reported interstate travel ban as a hindrance to the access of PLHIV to ART in some other countries [20, 51]. Data from the current study also suggest that participants’ consideration of the possibility of negative side effects of the COVID-19 vaccine on their health had led to their decision not to receive the vaccine. Such consideration seemed to be influenced by misleading information they received from others about the vaccine and their lack of knowledge about the effectiveness of the vaccine against COVID-19 infection. In addition, it can also be argued that these factors may have led to the distrust among the participants towards the COVID-19 vaccine and healthcare providers, which could have corroborated their decision not to get vaccinated [52-54]. Thus, information dissemination and education on the COVID-19 vaccine to PLHIV and general community members are necessary to enhance both their knowledge of health benefits and uptake of the vaccine [55, 56]. Negative economic or financial impacts of COVID-19 reflected in disruption of income-generating activities and reduced or loss of incomes which led to the ability to afford access to healthcare-related expenses were also apparent among the participants and considered as barriers that impeded their access to ART service. It is plausible to allude that such a condition could have long-term economic or financial impacts on already poor participants and their families who experienced the loss of income during the pandemic, which may also influence other aspects of their life at both individual and familial levels. These are in line with the findings of previous studies which have reported that economic or financial hardship is a barrier that hindered access of PLHIV to ART and other healthcare services in general [45, 57, 58]. Similarly, long-distance travel to the HIV clinic which increased transport costs was also reported as influencing participants’ access to ART service during the pandemic, which is consistent with previous findings reported elsewhere [20, 24, 58].

### Study limitations and strengths

This study has some limitations that need to be considered when interpreting its findings. The use of the snowball sampling technique to recruit the participants and the dissemination of study information sheets through healthcare facilities were the limitations of the study as these had led us to recruiting participants from the same networks as the current participants. These may have led to an incomplete overview of barriers that hampered the access of PLHIV to ART service during the COVID-19 pandemic. However, to our knowledge, there has not been much evidence globally and in the context of Indonesia about barriers to access to ART during the COVID-19 pandemic from the views and experiences of PLHIV. Thus, the current findings provide useful information for the government and healthcare systems and facilities in Belu district and other parts of Indonesia, as well as other similar settings globally to develop strategies to support the accessibility of ART service among PLHIV during the pandemic.

## Conclusions

This study presents the views and experiences of PLHIV about barriers to their access to ART service during the COVID-19 pandemic. The barriers included fear of contracting COVID-19 transmission, lockdown and lack of information about ART service delivery during the COVID-19 pandemic, mandatory COVID-19 vaccine certificate for inter-district travellers and financial barriers. The findings indicate the need for dissemination of information about the provision of ART service during the pandemic and the health benefits of the COVID-19 vaccine for PLHIV. The findings also indicate the need for new strategies to bring HIV care services, especially ART closer to PLHIV during the pandemic such as a community-based delivery strategy through the utilisation of existing networks of community health workers which have been proven effective in other countries [59] and coordination to provide ART at public health centres which are available in each sub-district. Future large-scale studies exploring views and experiences of PLHIV about barriers to their access to HIV care services during the COVID-19 pandemic are recommended.

## Data Availability

All data produced in the present work are contained in the manuscript

## List of Abbreviations

AIDS: Acquired Immune Deficiency Syndrome
ART: Antiretroviral Therapy
COVID-19: Coronavirus Disease
PLHIV: People Living with HIV
WHO: World Health Organisation

## Declarations

### Ethics approval and consent to participants

Ethics approval for this study was obtained from the Health Research Ethics Committee, Duta Wacana Christian University, Indonesia (No. 1005/C.16/FK). All participants signed and returned informed consent on the interview day prior to commencing the interview.

### Consent for publication

Not applicable.

## Acknowledgements

We would like to thank the participants who had spent their time to voluntarily take part in the interview and provided us with valuable information.

## Authors’ contribution

NKF was involved in conceptualisation, development of the methodology, conducting formal analysis and writing the original draft of the paper and reviewing and editing the paper critically for important intellectual content. HAG was involved in conceptualisation, development of the methodology, and reviewing and editing the paper critically for important intellectual content. ALS was involved in conceptualisation, development of the methodology, project administration and investigation. PRW was involved in conceptualisation, development of the methodology, and reviewing and editing the paper critically for important intellectual content.

## Availability of data and materials

The data on which the conclusions of the manuscript rely are available in the paper. All datasets cannot be shared publicly because of sensitive information regarding the interviewees (PLHIV) and the restriction set by the human research ethics committees. Data presented in this study are available on request from the corresponding author for researchers who meet the criteria for access to confidential data.

## Author details

^1^Research Centre for Public Health, Equity and Human Flourishing, Torrens University Australia, Adelaide, South Australia, Australia. ^2^Institute of Resource Governance and Social Change, Kupang, Indonesia. ^3^College of Health Sciences, Mekelle University, Mekelle, Tigray, Ethiopia. ^4^Atapupu Public Health Centre, Health Department of Belu District, Indonesia.

